# Neutrophil calprotectin identifies severe pulmonary disease in COVID-19

**DOI:** 10.1101/2020.05.06.20093070

**Authors:** Hui Shi, Yu Zuo, Srilakshmi Yalavarthi, Kelsey Gockman, Melanie Zuo, Jacqueline A. Madison, Christopher Blair, Wrenn Woodward, Sean P. Lezak, Njira L. Lugogo, Robert J. Woods, Christian Lood, Jason S. Knight, Yogendra Kanthi

## Abstract

Severe cases of coronavirus disease 2019 (**COVID-19**) are regularly complicated by respiratory failure. While it has been suggested that elevated levels of blood neutrophils associate with worsening oxygenation in COVID-19, it is unknown whether neutrophils are drivers of the thrombo-inflammatory storm or simple bystanders. To better understand the potential role of neutrophils in COVID-19, we measured levels of the neutrophil activation marker S100A8/A9 (calprotectin) in hospitalized patients and determined its relationship to severity of illness and respiratory status. Patients with COVID-19 (n=172) had markedly elevated levels of calprotectin in their blood. Calprotectin tracked with other acute phase reactants including C-reactive protein, ferritin, lactate dehydrogenase, and absolute neutrophil count, but was superior in identifying patients requiring mechanical ventilation. In longitudinal samples, calprotectin rose as oxygenation worsened. When tested on day 1 or 2 of hospitalization (n=94 patients), calprotectin levels were significantly higher in patients who progressed to severe COVID-19 requiring mechanical ventilation (8039 ± 7031 ng/ml, n=32) as compared to those who remained free of intubation (3365 ± 3146, p<0.0001). In summary, serum calprotectin levels track closely with current and future COVID-19 severity, implicating neutrophils as potential perpetuators of inflammation and respiratory compromise in COVID-19.

## INTRODUCTION

Since December 2019, the outbreak of coronavirus disease 2019 **(COVID-19)** caused by severe acute respiratory syndrome coronavirus 2 (**SARS-CoV-2**) has spread to hundreds of countries and territories and has been declared a global pandemic. Severe COVID-19 results in death due to progressive hypoxemia, acute respiratory distress syndrome (**ARDS**), and multi-organ failure [1]. The role of the host response in this progression remains to be fully defined.

S100A8 (myeloid-related protein 8/MRP8) and S100A9 (MRP14) are calcium-binding proteins that belong to the S100 family. They exist mainly together as a biologically-functional heterodimer known as S100A8/A9 or calprotectin. Calprotectin is found in abundance in neutrophils, where it can account for almost two-thirds of soluble protein in the cytosol. Calprotectin may also be detected at low levels in monocytes, macrophages, platelets, and squamous epithelial cells [2]. Upon neutrophil activation or death, calprotectin is released extracellularly where it has microbicidal functions (via heavy-metal chelation) and also serves as a pro-inflammatory ligand for innate receptors such as receptor for advanced glycation endproducts (RAGE) and Toll-like receptor 4 (TLR4) [3]. Given its small size, easy diffusion between tissue and blood, and resistance to enzymatic degradation, calprotectin is a sensitive and dynamic marker of neutrophil activation anywhere in the body [4, 5]. High levels of calprotectin have been found in many types of infectious and inflammatory diseases—including sepsis, myocardial infarction, inflammatory bowel disease, lupus, and adult-onset Still’s disease—where it tracks closely with disease severity [6-10].

While work to date exploring COVID-19 pathophysiology has focused especially on macrophages and their products such as interleukin-6 (IL-6) and IL-1β, it has also been observed that elevated levels of blood neutrophils associate with worsening oxygenation in COVID-19 [11-13]. Furthermore, our group and others have also revealed a potentially pathogenic role for neutrophil-derived extracellular traps (NETs) in COVID-19 [14, 15]. There remains though a paucity of information about neutrophil catalysts, checkpoints, and effector mechanisms in COVID-19—all of which could add actionable context to our understanding of the COVID-19 thrombo-inflammatory storm. Here, to better understand the potential role of neutrophils in COVID-19, we measured calprotectin in the blood of patients hospitalized with COVID-19 and determined its relationship to severity of illness and respiratory status.

## RESULTS AND DISCUSSION

Serum samples were obtained from 172 patients hospitalized with COVID-19 at a large academic hospital (**Supplemental Table 1**). In some cases, sera were stored at 4°C in the clinical laboratory for up to 48 hours before being frozen. Interestingly, we found that calprotectin levels are stable in both serum and plasma for up to six days at 4°C (**Supplemental Figure 1**), which is in line with past research on the topic [16]. As compared with serum samples from 47 healthy controls, the COVID-19 samples showed markedly higher levels of calprotectin (**Figure 1A**). For 36 patients, longitudinal sera were available. Nine of those patients showed a clinically meaningful change in oxygenation status during the period of collection (six worsening and three improving). Notably, calprotectin levels trended upward in the six patients for whom oxygenation worsened (**Figure 1B**). We next asked how calprotectin compared to commonly available clinical measurements. Specifically, we assessed potential correlations with C-reactive protein, ferritin, lactate dehydrogenase, absolute neutrophil count, absolute lymphocyte count, hemoglobin level, and platelet count. Calprotectin demonstrated a positive correlation with C-reactive protein (**Figure 1C**), ferritin (r=0.31, p=0.0002), lactate dehydrogenase (r=0.52, p<0.0001), absolute neutrophil count (**Figure 1D**), and platelet count (r=0.39, p<0.0001). There was no correlation with absolute lymphocyte count (**Figure 1E**), and a negative correlation with hemoglobin level (r=-0.34, p<0.0001). In summary, calprotectin is markedly elevated in the sera of patients with COVID-19 and may rise as clinical status deteriorates.

**Figure 1:**
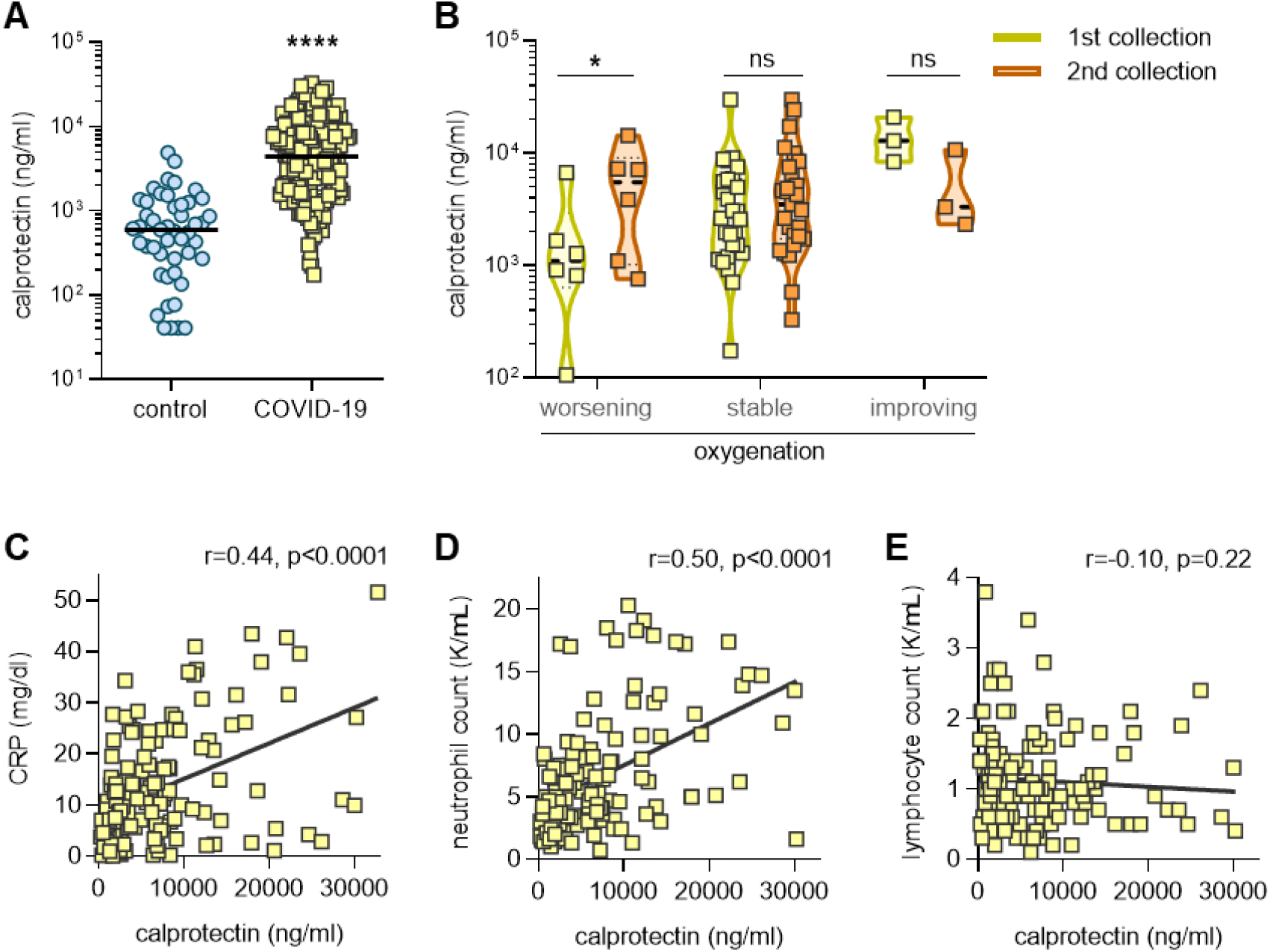
Calprotectin in sera of COVID-19 patients and its association with clinical studies. **A**, Sera from COVID-19 patients (n=172) and healthy controls (n=47) were assessed for calprotectin (note log scale). COVID-19 samples were compared to controls by Mann-Whitney test; ****p<0.0001. **B**, For 36 patients, serum samples from two time points were available. Patients were grouped by whether their oxygenation was worsening, stable, or improving; *p<0.05 by paired Wilcoxon test. **C-E**, Calprotectin levels were compared to clinical laboratory results (when available on the same day as the research sample). Spearman’s correlation coefficients were calculated for C-reactive protein (n=138), absolute neutrophil count (n=139), and absolute lymphocyte count (n=139).

We next determined each patient’s clinical respiratory status at the time calprotectin was measured. As compared with patients breathing room air, patients requiring mechanical ventilation had significantly higher levels of calprotectin (**Figure 2A**). Interestingly, differences were also appreciated between patients requiring noninvasive oxygen support (such as nasal-cannula oxygen) and mechanical ventilation (**Figure 2A**). In contrast, C-reactive protein did not discriminate between patients requiring noninvasive oxygen support and those requiring mechanical ventilation (**Figure 2B**). To further evaluate the potential clinical utility of calprotectin, we performed receiver operating characteristic (ROC) curve analysis based on requirement for mechanical ventilation. As compared with C-reactive protein, ferritin, and lactate dehydrogenase, calprotectin had a superior area under the curve (**Figure 2C**). Beyond clinical respiratory status, oxygenation efficiency can also be measured by comparing pulse oximetry (SpO2) to the fraction of inspired oxygen (FiO2). We tested the correlation between calprotectin and SpO2/FiO2 ratio, and found a striking negative association (**Figure 2D**). A less robust association was also appreciated for C-reactive protein (**Figure 2E**). In summary, calprotectin levels strongly associate with severe respiratory disease requiring mechanical ventilation.

**Figure 2:**
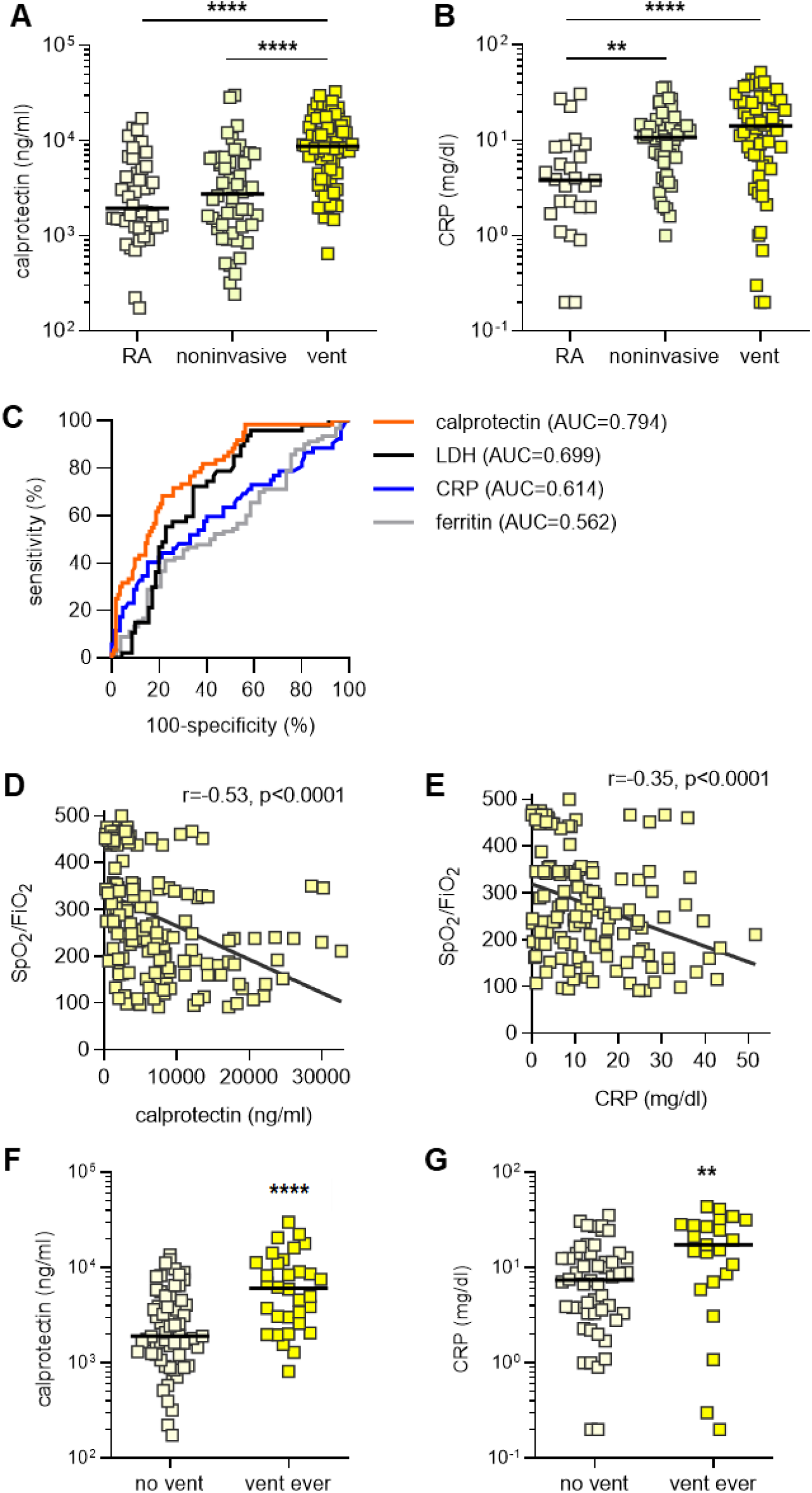
Levels of calprotectin track closely with oxygenation status. **A-B**, Patients (n=172) were grouped by clinical status: room air (n=41), noninvasive supplemental oxygen (n=71), or mechanical ventilation (n=60). Levels of calprotectin and C-reactive protein were compared by Kruskal-Wallis test corrected by Dunn’s test for multiple comparisons; *p<0.05 and ****p<0.0001. **C**, Receiver operating characteristic curves based on requirement for mechanical ventilation. **D-E**, Calprotectin (n=172) or C-reactive protein (n=137) were compared to SpO2/FiO2 ratio for each patient, and correlations were determined by Spearman’s test. **F-G**, For 94 patients, a calprotectin level was available from hospital day 1 or 2. For 75 of the 94, a CRP level was also available. Patient were then grouped by whether they at any point required mechanical ventilation (vent ever, n=32) during their hospitalization. Groups were compared by Mann-Whitney test; **p<0.01 and ****p<0.0001.

To confirm these findings, we also obtained plasma from 119 of the 172 patients. As compared with plasma samples from 50 healthy controls, the COVID-19 samples showed markedly higher levels of calprotectin (**Supplemental Figure 2A**). Furthermore, for the COVID-19 patients, plasma calprotectin demonstrated a distinct negative correlation with SpO2/FiO2 ratio (**Supplemental Figure 2B**), and positive correlations with C-reactive protein (**Supplemental Figure 2C**) and absolute neutrophil count (**Supplemental Figure 2D**). We also found positive correlations between calprotectin and markers of neutrophil extracellular trap (NET) release including cell-free DNA (**Supplemental Figure 2E**) and myeloperoxidase-DNA complexes (**Supplemental Figure 2F**).

Finally, of the 172 patients with serum samples evaluated here, 94 had sera available from the first two days of their hospitalization. When some of the aforementioned correlation analyses were reanalyzed with just these 94 samples (**Supplemental Figure 3**), we found a strong negative correlation with SpO2/FiO2, and strong positive correlations with C-reactive protein and absolute neutrophil count. Most importantly, calprotectin levels were significantly higher in those individuals who required mechanical ventilation at any point during their hospitalization (n=32), as compared with those who did not (p<0.0001, **Figure 2F**). C-reactive protein was also analyzed (when available on the same day as the calprotectin measurement) and was found to be predictive of any mechanical ventilation with a p-value of 0.0054 (**Figure 2G**). Taken together, these data suggest a compelling relationship between neutrophil activation, as defined by serum calprotectin levels, and severe respiratory disease in COVID-19.

In summary, we report markedly elevated levels of serum and plasma calprotectin in the majority of patients hospitalized with COVID-19. Furthermore, we found that high levels of serum calprotectin on day 1 or 2 of hospitalization tracked with a requirement for mechanical ventilation at any point during the admission, a finding that should be assessed in larger prospective cohorts. These data provide strong evidence in support of neutrophils as potential players in moderate-to-severe cases of COVID-19.

Our study raises the need to investigate the specific form of neutrophil activation and/or cell death that floods COVID-19 blood with excess calprotectin. Tissue damage and neutrophil necrosis are potential sources of passive calprotectin release [17]. At the same, active calprotectin secretion has been documented upon stimulation of neutrophils with complement C5 and fMLP [18]. Engagement of neutrophil PSGL-1 by E-selectin can also trigger neutrophils to actively release calprotectin in calcium ion-dependent fashion (TLR4 and its downstream MyD88 and Rap1-GTP trigger release via this same mechanism) [19]. A newer consideration regarding active calprotectin release is NETosis [14, 20]. NETs are extracellular webs of DNA, histones, and microbicidal proteins that appear to perpetuate many types of lung disease including smoking-related disease, cystic fibrosis, and ARDS. NETs leverage calprotectin as an antimicrobial strategy against Candida [21] and Aspergillus [22], but, when left unchecked, NETs are also an important source of macrophage activation and microvascular occlusion. Whether passive calprotectin release in necrotic lung tissue or active release via NETosis (or another mechanism) is most important to COVID-19 pathophysiology awaits further research.

In addition to being an inflammatory marker, calprotectin may also have a direct role in the self-amplifying thrombo-inflammatory storm of COVID-19 via engagement and activation of innate immune sensors such as RAGE [23, 24] and TLR4 [25, 26]. Depending on the system, calprotectin has also been detected both upstream [27] and downstream [28] of IL-6, which has emerged as a possible therapeutic target in COVID-19. As a key alarmin molecule of the immune system, calprotectin modulates the inflammatory response by recruiting leukocytes and stimulating cytokine secretion [29]. Calprotectin also induces reactive nitrogen and oxygen species [30, 31] and triggers microvascular endothelial cells to take on thrombogenic and pro-inflammatory phenotypes characterized by increased vascular permeability and synthesis of cytokines, chemokines, and adhesion molecules [29]. Furthermore, calprotectin is a potent stimulator of neutrophils themselves, promoting degranulation and phagocytosis [32, 33], as well as NETosis [34]. Intriguingly, crosstalk between neutrophils, platelets, and calprotectin appears to play a role in both arterial and venous thrombosis [34, 35], which are being increasingly identified as complications of COVID-19 [36, 37].

As we await definitive antiviral and immunologic solutions to the current pandemic, we posit that anti-neutrophil therapies [38-40] may be part of a personalized strategy for some individuals affected by COVID-19 who are at risk for progression to respiratory failure. In this context, calprotectin is well-positioned to be an early indicator of patients with COVID-19 likely to progress to respiratory failure and who therefore require immunomodulatory treatment.

## Data Availability

Once the manuscript has been published in a peer-reviewed journal, the corresponding author will make all data available upon request.

## AUTHORSHIP

HS, YZ, SY, KG, MZ, JM, CNB, WW, and SPL conducted experiments and analyzed data. HS, YZ, NLL, RJW, CL, JSK, and YK conceived the study and analyzed data. All authors participated in writing the manuscript and gave approval before submission.

## ACKNOWLEDGEMENTS

The work was supported by a COVID-19 Cardiovascular Impact Research Ignitor Grant from the Michigan Medicine Frankel Cardiovascular Center as well as by the A. Alfred Taubman Medical Research Institute. YZ was supported by career development grants from the Rheumatology Research Foundation and APS ACTION. JAM was partially supported by the VA Healthcare System. JSK was supported in part by grants from the NIH (R01HL115138), Lupus Research Alliance, and Burroughs Wellcome Fund. YK was supported by part by the NIH (K08HL131993, R01HL150392), Falk Medical Research Trust Catalyst Award, the JOBST-American Venous Forum Award, the Intramural Research Program of the NIH and NHLBI, and the Lasker Foundation.

## SUPPLEMENTAL MATERIAL

### EXTENDED METHODS

#### Human samples

Serum and plasma samples from 172 hospitalized COVID-19 patients were used in this study. Blood was collected into serum separator tubes or EDTA tubes by a trained hospital phlebotomist. After completion of biochemical and hematological testing ordered by the clinician, the remaining serum and plasma was stored at 4°C for up to 48 hours before it was released to the research laboratory. Serum and plasma samples were immediately divided into small aliquots and stored at −80°C until the time of testing. All 172 patients had a confirmed COVID-19 diagnosis based on FDA-approved RNA testing. This study complied with all relevant ethical regulations, and was approved by the University of Michigan Institutional Review Board (HUM00179409), which waived the requirement for informed consent given the discarded nature of the samples. Healthy volunteers were recruited through a posted flyer; exclusion criteria for these controls included history of a systemic autoimmune disease, active infection, and pregnancy. For serum analysis, 47 controls were used (36 females and 11 males with mean age of 38.8 ± 11.3). For plasma analysis, 50 controls were used (37 females and 13 males with mean age of 41.5 ± 13.4).

#### Quantification of S100A8/A9 (calprotectin)

Calprotectin levels were measured with the Human S100A8/S100A9 Heterodimer DuoSet ELISA (DY8226-05, R&D Systems) according to the manufacturer’s instructions.

#### Quantification of cell-free DNA

Cell-free DNA was quantified in sera using the Quant-iT PicoGreen dsDNA Assay Kit (Invitrogen, P11496) according to the manufacturer’s instructions.

#### Quantification of myeloperoxidase-DNA complexes

Myeloperoxidase-DNA complexes were quantified similarly to what has been previously described. This protocol used several reagents from the Cell Death Detection ELISA kit (Roche). First, a high-binding EIA/RIA 96-well plate (Costar) was coated overnight at 4°C with anti-human myeloperoxidase antibody (Bio-Rad 0400-0002), diluted to a concentration of 1 µg/ml in coating buffer (Cell Death kit). The plate was washed two times with wash buffer (0.05% Tween 20 in PBS), and then blocked with 4% bovine serum albumin in PBS (supplemented with 0.05% Tween 20) for 2 hours at room temperature. The plate was again washed five times, before incubating for 90 minutes at room temperature with 10% serum or plasma in the aforementioned blocking buffer (without Tween 20). The plate was washed five times, and then incubated for 90 minutes at room temperature with 10x anti-DNA antibody (HRP-conjugated; from the Cell Death kit) diluted 1:100 in blocking buffer. After five more washes, the plate was developed with 3,3’,5,5’-Tetramethylbenzidine (TMB) substrate (Invitrogen) followed by a 2N sulfuric acid stop solution. Absorbance was measured at a wavelength of 450 nm using a Cytation 5 Cell Imaging Multi-Mode Reader (BioTek). Data were normalized to *in vitro*-prepared NET standards included on every plate, which were quantified based on their total protein content.

#### Statistical analysis

When two groups were present, normally-distributed data were analyzed by two-sided t test and skewed data were analyzed by Mann-Whitney test or Wilcoxon test. For three or more groups, analysis was by one-way ANOVA or Kruskal-Wallis test with correction for multiple comparisons. Correlations were tested by Spearman’s method. Data analysis was with GraphPad Prism software version 8. Statistical significance was defined as p<0.05.

---

**Supplemental Table 1:**
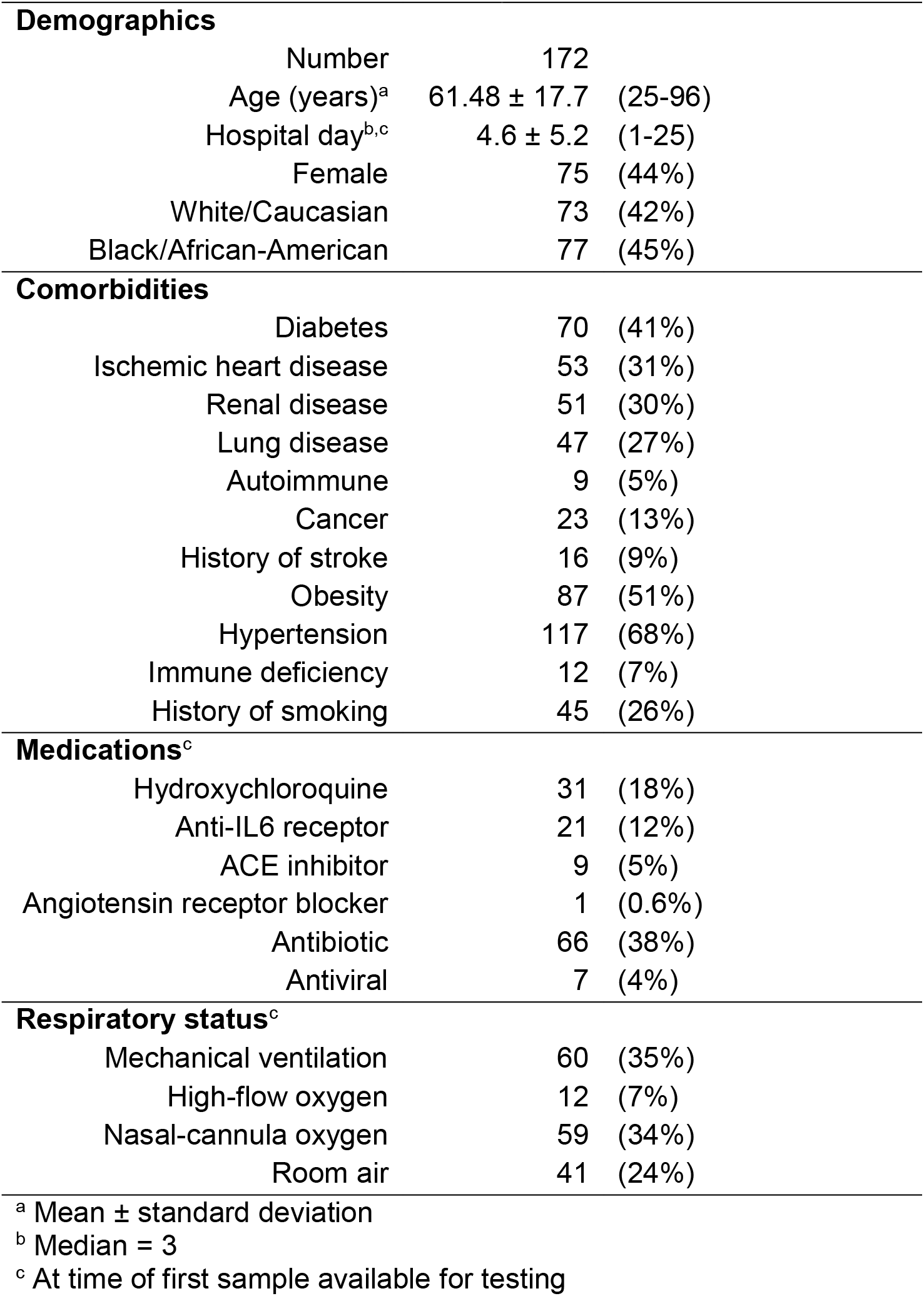
COVID-19 patient characteristics.

**Supplemental Figure 1:**
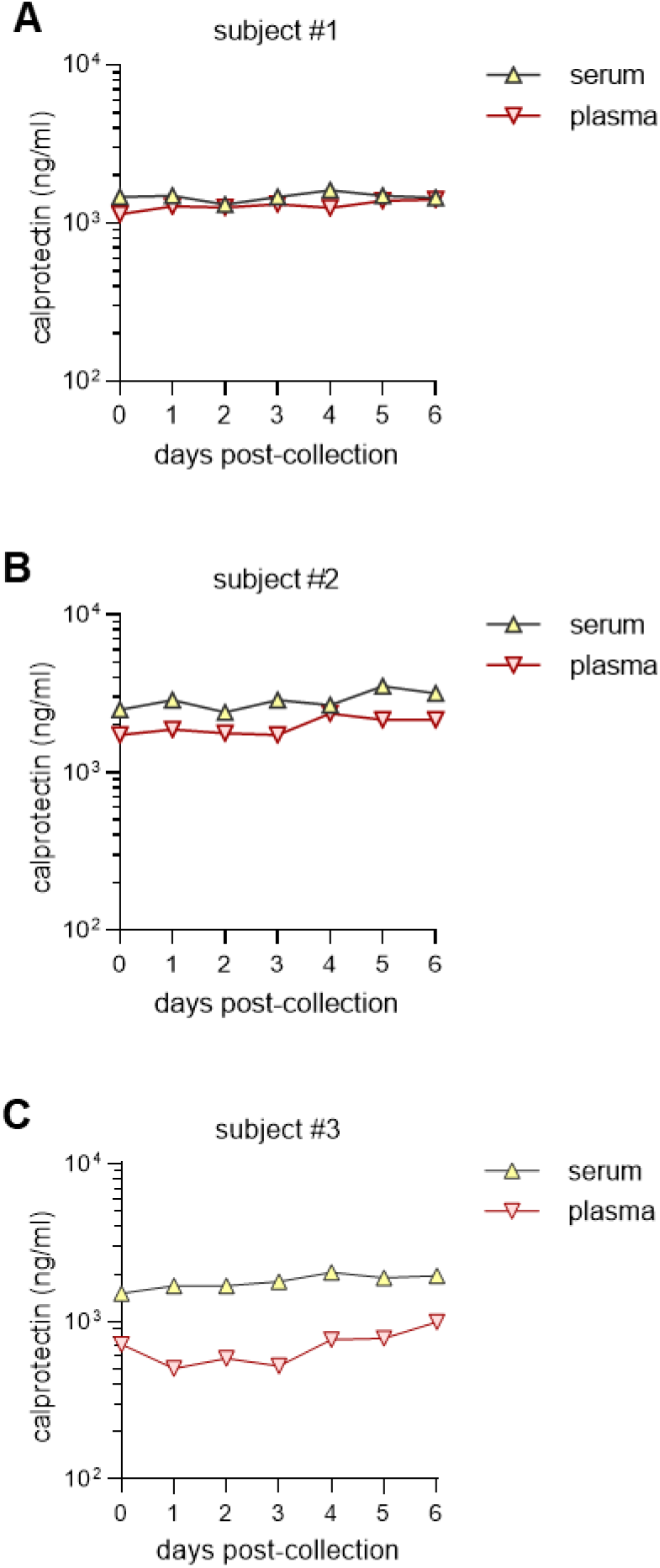
Stability of calprotectin in serum and plasma. Serum and plasma were prepared on day=0 from three healthy individuals. The samples were stored at 4°C with an aliquot removed daily for freezing at −80°C. After 7 aliquots had been frozen, samples were thawed and calprotectin levels were determined.

**Supplemental Figure 2:**
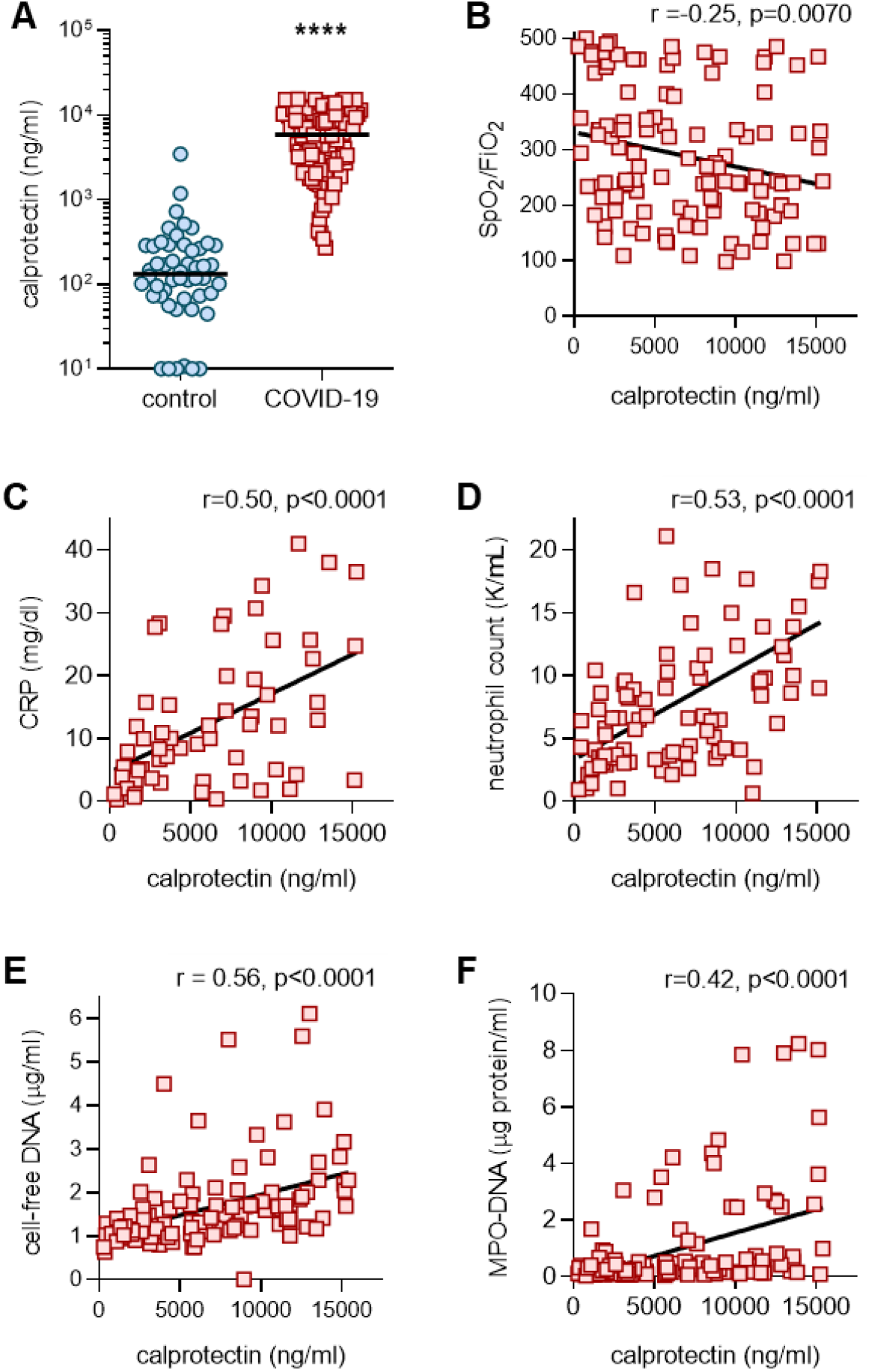
Calprotectin in plasma of COVID-19 patients. **A**, Plasma from COVID-19 patients (n=119) and healthy controls (n=50) were assessed for calprotectin. COVID-19 samples were compared to controls by Mann-Whitney test; ****p<0.0001. **B**, Calprotectin was compared to SpO2/FiO2 ratio for each patient (n=115), and correlation was determined by Spearman’s test. **C-F**, Calprotectin levels were compared to clinical laboratory results (when available on the same day as the research sample) or markers of neutrophil extracellular traps (NETs). Spearman’s correlation coefficients were calculated for C-reactive protein (n=59), absolute neutrophil count (n=87), cell-free DNA (n=118), and myeloperoxidase-DNA complexes (n=119).

**Supplemental Figure 3:**
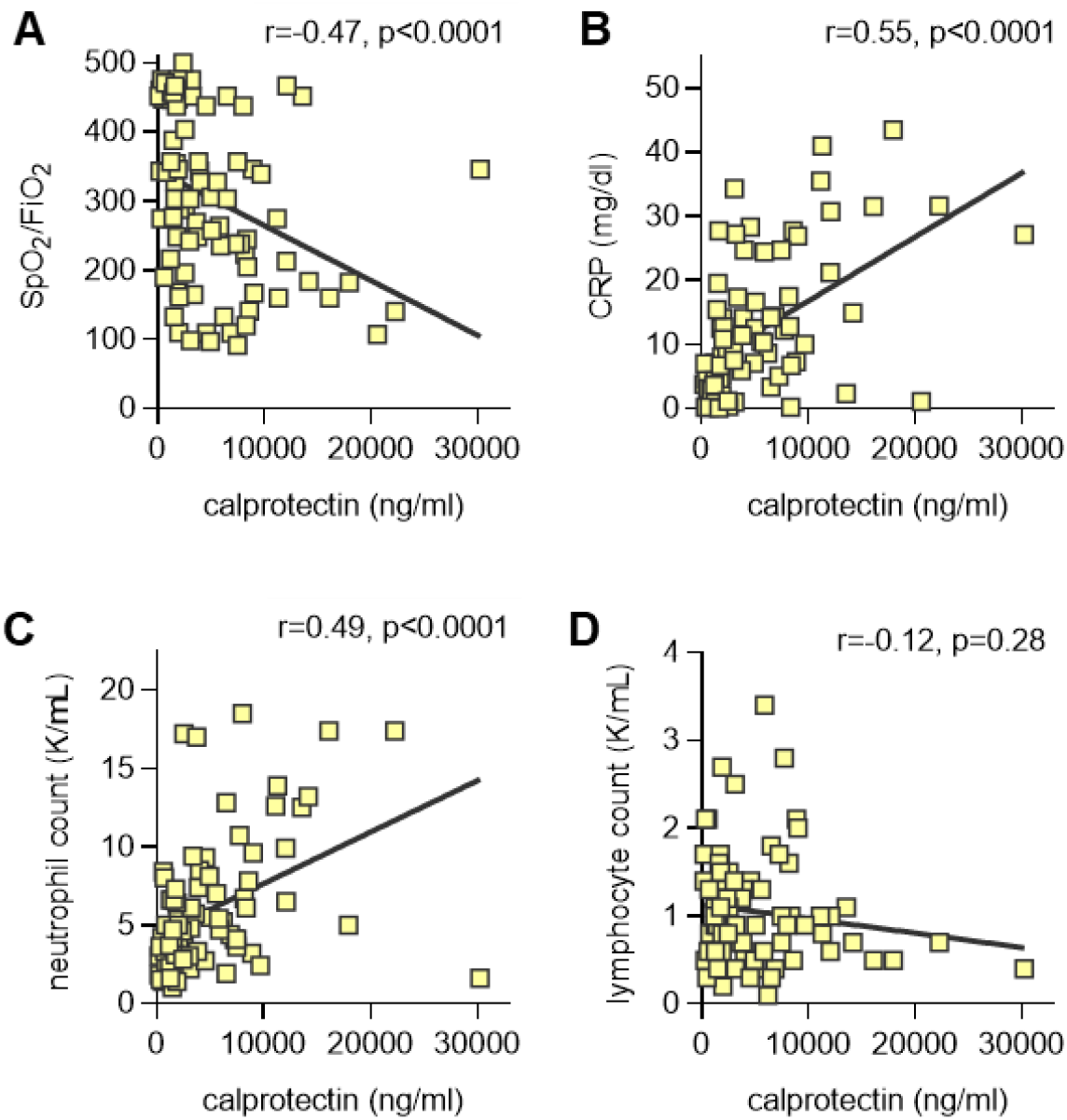
Correlation analysis for the 94 patients who had samples available on day 1 or 2 of hospitalization. **A-D**, Calprotectin levels were compared to oxygenation efficiency or clinical laboratory results (when available on the same day as the research sample). Spearman’s correlation coefficients were calculated for SpO2/FiO2 ratio (n=93), C-reactive protein (n=79), absolute neutrophil count (n=82), and absolute lymphocyte count (n=82).

